# Systematic comparison of observational and Mendelian Randomization estimates for cardiometabolic proteomic signatures

**DOI:** 10.64898/2025.12.02.25341031

**Authors:** Thorarinn Jonmundsson, Valur Emilsson, Elisabet A. Frick, Heida Bjarnadottir, Eva Jacobsen, Thor Aspelund, Lenore J. Launer, Joseph J. Loureiro, Anthony P. Orth, Nancy Finkel, Vilmundur Gudnason, Valborg Gudmundsdottir

## Abstract

Proteomic Mendelian randomization (MR) studies have gained widespread interest due to their potential to reveal novel therapeutic targets. Discrepancies are often reported between observational and genetic estimates of protein-trait associations, but the factors influencing directional agreement remain poorly understood. We systematically evaluated overlaps and directional agreement between observational associations and bi-directional two-sample MR estimates for 7,288 serum protein measurements across 25 cardiometabolic traits in the AGES-RS cohort (n = 5,364). Overall, our findings demonstrate that the proteomic signatures of cardiometabolic traits appear to be predominantly shaped by reverse causation, as observational associations significantly overlapped with those from the reverse (protein → phenotype) MR and showed nearly uniform directional agreement (median 100%). By contrast, observational signatures did generally not enrich for forward (phenotype → protein) MR associations and showed only weak directional agreement (median 55%), suggesting the two approaches largely capture orthogonal biological pathways where causal signals potentially reflect the effects of widespread molecular pleiotropy. Restricting causal analyses to observationally significant proteins proved both limiting and redundant, disproportionately enriching for reverse and ambiguous causal associations. Coding variant burden was the strongest positive predictor of directional agreement for forward MR estimates, possibly reflecting their direct effects on protein structure and function. Although forward MR remains a valuable approach for prioritizing causal candidates, our findings caution against interpreting circulating levels as direct causal exposures.

## Introduction

Recent advances in protein quantification technologies have enabled large-scale proteome-wide studies in human cohorts, offering unprecedented insights into the human proteome^1,2^. Among these, genome-wide association studies (GWAS) of the circulating proteome have become a key tool for linking genetic variation to protein levels, laying the foundation for proteomic Mendelian randomization (MR) studies that aim to infer causal effects of proteins on complex traits^1,3,4^. Such proteomic MR studies have gained widespread interest due to their potential to reveal novel therapeutic targets^5,6^.

A common approach in proteomic cohort studies is to identify protein predictors of an outcome of interest, and then evaluate those candidates for causal effects using MR^7^. Unlike observational studies, which estimate direct associations between measured protein levels and phenotypic outcomes, MR uses genetic instruments to mitigate confounding and reverse causation^8,9^. While not all proteomic MR studies compare MR estimates to observational associations^10–12^, doing so provides an opportunity to evaluate the consistency of directional effects between these two approaches. This consistency can be assessed by calculating the ratio of agreement, defined as the proportion of directionally concordant protein-phenotype pairs for a given phenotype or group of interest. Somewhat surprisingly, multiple proteomic MR studies have reported low directional agreement between observational and MR estimates across protein quantification platforms, phenotypes, and cohorts. For example, Titova et al.^13^ reported 43% agreement for incident myocardial infarction using the Olink platform, Schuermans et al.^14^ found 54% agreement for coronary artery disease and heart failure in UK Biobank, and Gudmundsdottir et al.^15^ observed 56% agreement for type 2 diabetes in the Age, Gene/Environment Susceptibility–Reykjavik Study (AGES-RS) using the SomaScan 5k assay.

These recurring low-agreement estimates point to a disconnect between genetically determined and measured circulating protein levels in their associations with disease. They also raise several questions: What is the observed directional agreement when evaluated systematically across traits? Do phenotype identity and instrument composition influence deviation in directional agreement? And to what extent does reverse causation shape proteomic signatures in disease contexts? To address these questions, we systematically evaluated thedirectional agreement between observational and bi-directional MR associations for 7,288 SOMAmers targeting 6,385 serum proteins across 25 cardiometabolic phenotypes in the AGES-RS cohort^16^ using the SomaScan 7K (v4.1) platform^17^. We compared results with and without filtering on proteins with observational significance and developed a hierarchical Bayesian model to quantify directional agreement, accounting for phenotype-specific effects, phenotype groupings, instrument strength, and coding variant burden. These features were chosen for their relevance to MR: instrument strength is a known determinant of estimate precision^18^, while coding variant burden may reflect stronger or more direct links between genetic variants and protein function^2^.

Our study reveals a global pattern across phenotypes, where observational proteomic signatures overlap significantly with reverse MR findings (the influence of disease liability on circulating protein levels) and are consistently directionally concordant, while forward MR estimates (the influence of genetically predicted circulating protein levels on disease risk) are consistently discordant. We highlight coding variant burden as a key driver of concordance, and demonstrate how observational filtering, instrument characteristics, and biological context influence causal inference in proteomic MR.

## Results

### The observed proteomic signatures of cardiometabolic traits

The characteristics of the AGES-RS cohort are summarized in Table 1, including the 25 cardiometabolic traits considered in this study. Out of 7,288 SOMAmers analyzed (Fig. 1), the vast majority or 6,400 (88%) contributed to the observational proteomic signature of at least one cardiometabolic phenotype, defined as SOMAmer–phenotype associations reaching a phenotype-specific Bonferroni threshold (P < 0.05/7,288) (Supplementary Table 1). These 6,400 SOMAmers accounted for a total of 36,242 SOMAmer–phenotype association pairs across 25 cardiometabolic traits. Of these pairs, 29,359 (81%) also met a study-wide threshold (P < 0.05/7288/25). Given the stability of the signatures across thresholds, all phenotype-specific associations were retained for subsequent analyses (Figure 2A; Supplementary Table 1). The median size of cardiometabolic proteomic signatures was 873 SOMAmer associations (IQR: 136-2,429), ranging from 2 associations for incident stroke to 4,124 for eGFR (Supplementary Table 2). Risk factors yielded more significant SOMAmer-phenotype pairs than disease phenotypes (median [IQR]: 2,356 [1,778–2,696] vs. 249 [83–779]). Among disease phenotypes, a greater number of significant associations were observed for prevalent than for incident disease (median [IQR]: 810 [349–1,358] vs. 102 [74–241]) (Figure 2A; Supplementary Table 2).

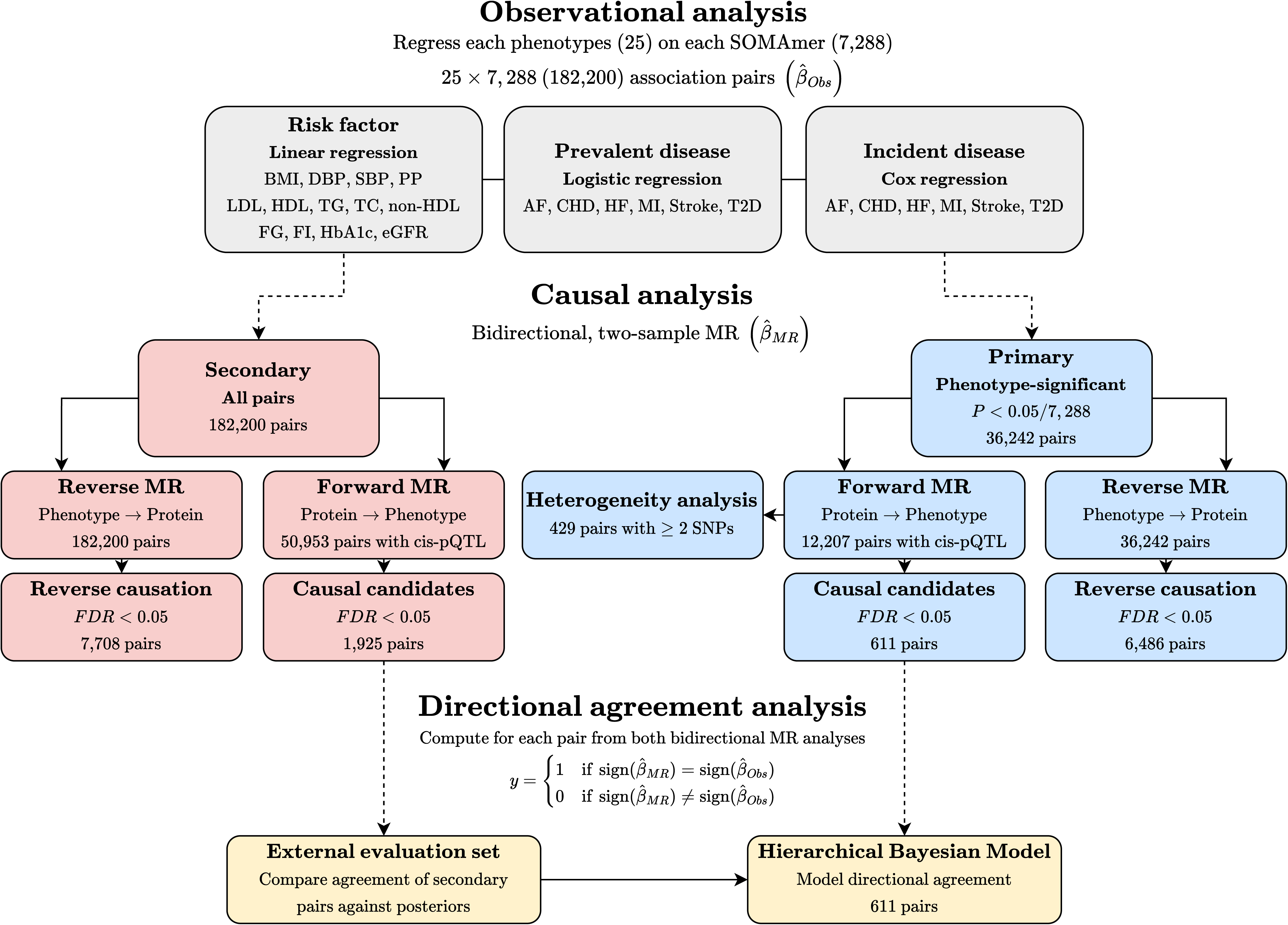

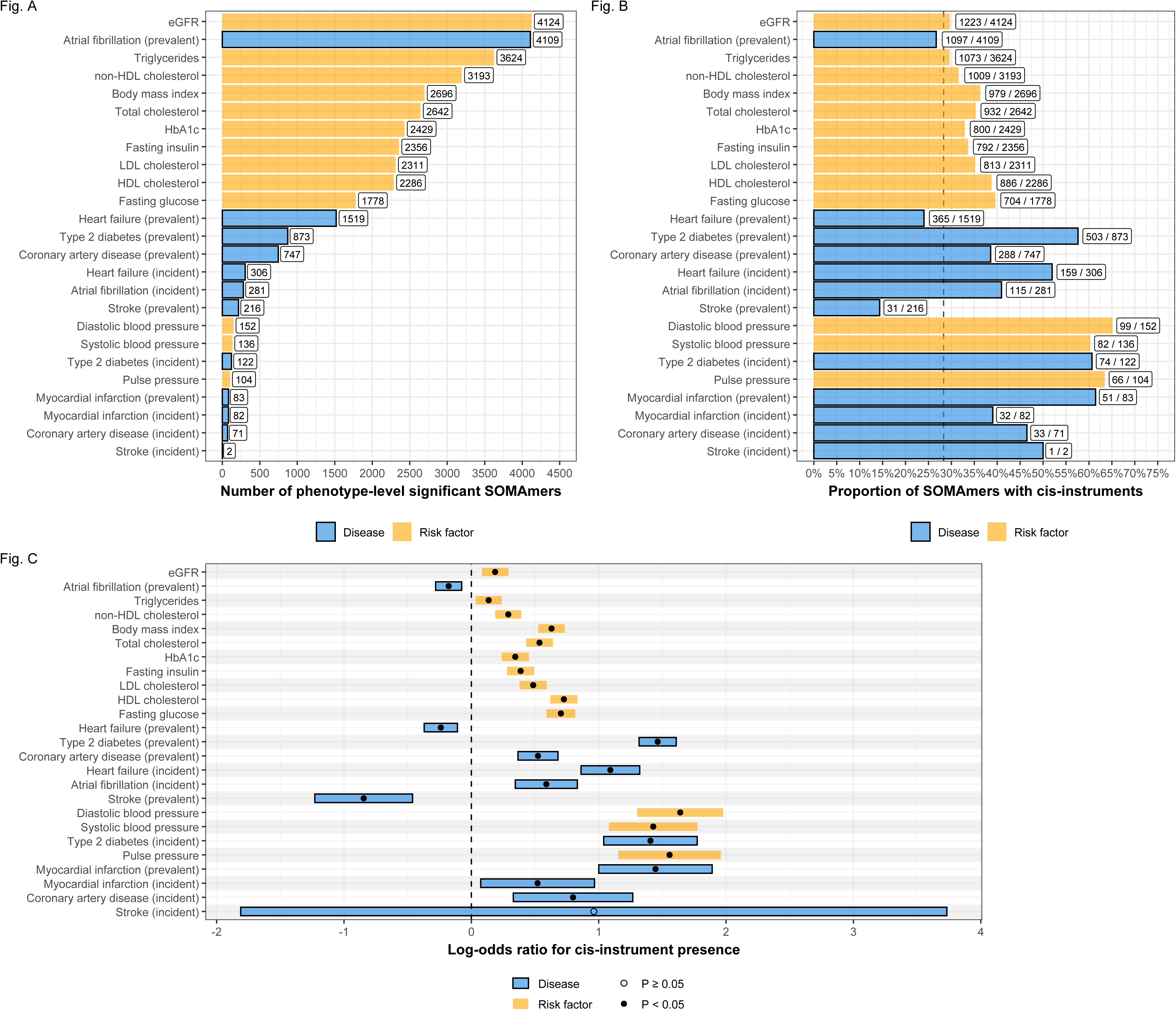

### Observational proteomic signatures are enriched for proteins with cis-instruments

Of 7,288 SOMAmers analyzed, 2,062 (28%) had at least one cis-acting instrument, a prerequisite to be tested in a forward MR framework. Across the cardiometabolic phenotypes, these corresponded to 50,953 cis-instrumented SOMAmer-phenotype pairs, i.e. the forward MR eligibility set, which includes pairs regardless of observational significance (Supplementary Table 3). Among the 36,242 pairs forming the observational proteomic signatures, 12,207 (34%) overlapped with this eligibility set (Figure 2B; Supplementary Table 3). Overall, the proteomic signatures of cardiometabolic phenotypes tended to be enriched for cis-instrumented SOMAmers (Fig. 2B; Supplementary Table 4; Supplementary Fig. 2B). Median cis-coverage across phenotypes was 39% (IQR: [33%-52%]), exceeding the expected 28% proportion under random distribution. Coverage was highest for DBP (65%), T2D (61%), and prevalent MI (61%), and lowest for prevalent stroke (14%), HF (24%), and AF (27%). When cis-protein enrichment was formally tested with logistic regression, 24 of the 25 phenotypes had significant log-odds ratios (P < 0.05) (Fig. 2C; Supplementary Table 5). Enrichment (positive log-odds ratios) was observed for 21 phenotypes, while prevalent stroke, HF, and AF showed significant depletion (negative log-odds ratio). The only non-significant phenotype was incident stroke, but that estimate was based on only two eligible pairs.

### Causal, and reverse causal, proteomic associations with cardiometabolic traits

Bidirectional two-sample MR analyses were first performed for the 36,242 SOMAmer-phenotype pairs from the observational proteomic signatures, using summary statistics from eleven external GWAS (Supplementary Table 6). In the forward MR analysis, 611 (5%) of 12,207 eligible pairs were identified as causal candidates (FDR < 0.05), corresponding to 350 unique SOMAmers across 21 phenotypes (Supplementary Table 7). The largest number of causal candidates was observed for TG (113 SOMAmers), whereas incident HF, incident MI, prevalent HF, and FI each yielded only one association (Figure 3A; Supplementary Table 7). Among the 611 causal candidates, 429 (70%) had more than one cis-pQTL and could be evaluated for instrument heterogeneity, of which 330 (77%) passed (Supplementary Table 8). Nearly all heterogeneous pairs belonged to risk factors (97 of 99 pairs), with the remaining two belonging to prevalent AF.

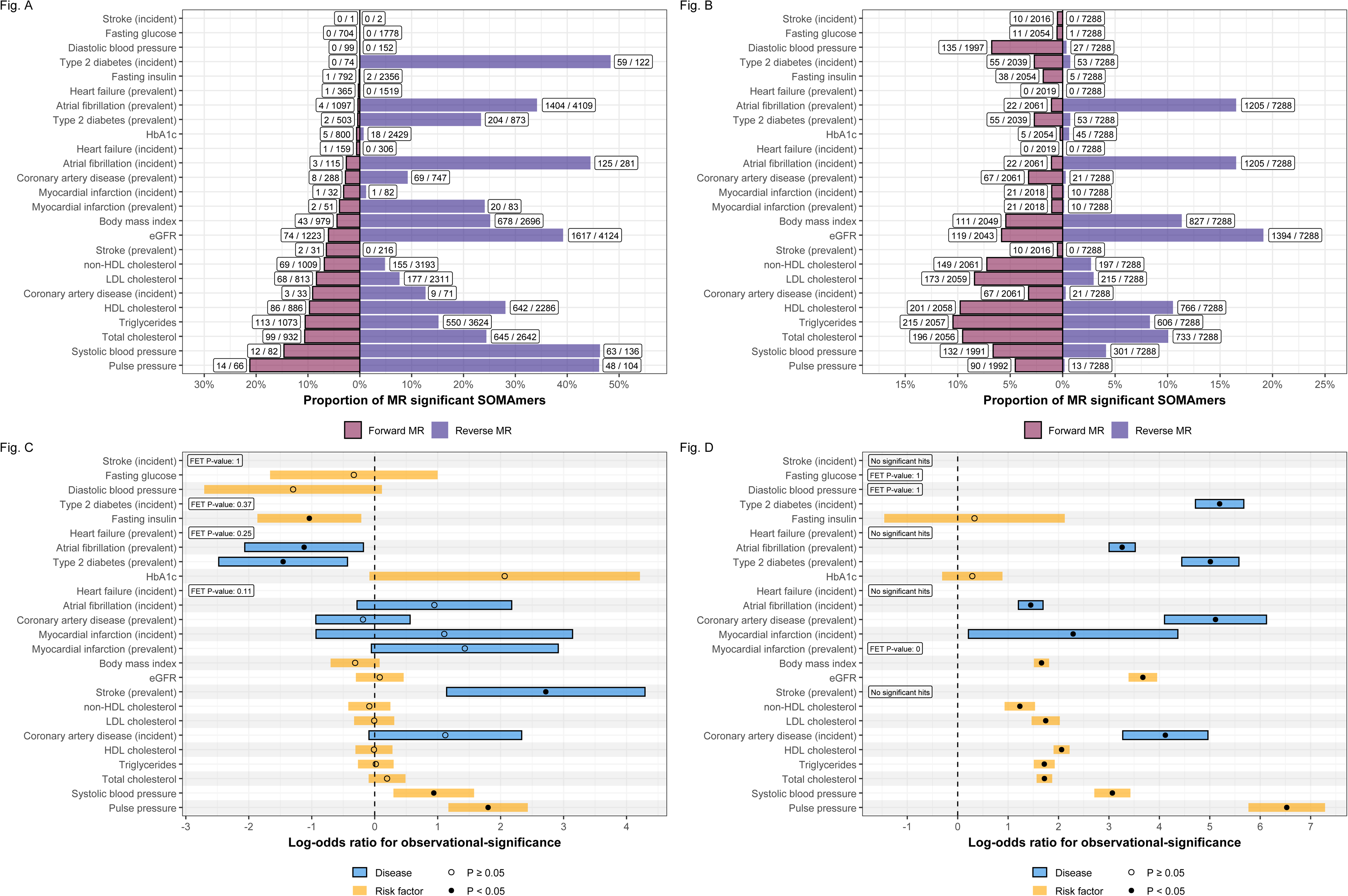

In the reverse MR analysis, 6,486 (18%) SOMAmer–phenotype pairs were significant (FDR < 0.05), spanning 19 phenotypes and 3,633 unique SOMAmers (Figure 3A; Supplementary Table 9). The strongest signal was observed for eGFR (1,617 SOMAmers), followed by prevalent AF (1,404), BMI (678 SOMAmers), and total cholesterol (645 SOMAmers). As in the forward analysis, incident MI and FI showed the fewest significant associations (one and two, respectively). No significant associations were detected for incident stroke, incident T2D, DBP, or FI in either direction. Likewise, the reverse analysis yielded no significant associations for either incident or prevalent HF or for prevalent stroke. Of the 611 forward-significant pairs, 188 (31%) were also significant in the reverse direction (Supplementary Table 10), reflecting proteins potentially affected by parallel mechanisms or feedback loops.

The secondary MR analysis included all SOMAmer-phenotype pairs, regardless of their observational significance. In the forward direction, 1,925 (4%) of the 50,953 eligible pairs were significant (FDR < 0.05), representing 23 phenotypes and 696 unique SOMAmers (Figure 3B; Supplementary Table 11). Notably, 595 (97%) of the 611 forward-significant pairs identified in the primary analysis were recovered in this more expanded analysis. In the reverse direction, 7,708 SOMAmer-phenotype pairs were significant (FDR < 0.05), spanning 21 phenotypes and 3,709 unique SOMAmers (Figure 3B; Supplementary Table 12). Consistent with the primary analysis, several overlaps were observed between directions. Of the 1,925 forward-significant pairs in the secondary analysis, 193 (10%) were also significant in the reverse direction (Supplementary Table 12); of those, 146 (76%) were also bidirectionally significant in the primary analysis (Supplementary Table 13).

### Observational proteomic signatures are enriched for reverse causation

We next assessed whether observationally significant SOMAmer-phenotype pairs were enriched for proteins with causal support from the MR analysis. For 76% (19/25) of the phenotypes, there was no association (logistic regression, P>0.05) between a protein’s observational significance and the probability of forward-MR significance (Fig. 3C; Supplementary Table 14). For three traits, observational significance was associated with greater probability of forward-MR significance: prevalent stroke (log-odds ratio [LOR] = 2.7; 95% CI [1.1; 4.3]), PP (LOR = 1.8; 95% CI [1.2; 2.4]) and SBP (LOR = 0.94; 95% CI [0.30; 1.6]). Conversely, significant depletion was observed for FI (LOR = -1.0; 95% CI [-1.9; -0.21]), prevalent AF (LOR = -1.1; 95% CI [-2.1; -0.18]), and prevalent T2D (LOR = -1.5; 95% CI [-2.5; - 0.43]). In the reverse direction, the enrichment pattern differed markedly (Fig. 3D; Supplementary Table 15). Observational significance was generally associated with higher probabilities of reverse-MR significance: 16 (76%) of 21 phenotypes that could be tested showed significant enrichment (P < 0.05) with positive LOR.

### Observed directional agreement between observational and MR estimates

All 611 SOMAmer–phenotype pairs identified in the primary forward MR analysis were assessed for directional agreement with observational estimates (Figure 4; Supplementary Tables 16-17). The overall agreement ratio was 55% (335 of 611 pairs), 54% (316 of 584 pairs) for risk factors, 70% (19 of 27 pairs) for diseases, 74% (14 of 19 pairs) for prevalent diseases, and 63% (5 of 8 pairs) for incident diseases (Supplementary Table 16). Agreement ratios were higher for prevalent and incident diseases than for risk factors, but these estimates were typically based on fewer causal candidates (often one to eight, compared with a median [IQR] of 68 [13-80] for risk factors). Among diseases, prevalent CAD showed particularly high agreement (88%; 7 of 8 pairs), whereas among risk factors, HDL-cholesterol had the highest agreement (67%; 58 of 86 pairs). Heterogeneity did not affect agreement (Fisher exact P value: 0.81) (Supplementary Table 8).

In the secondary forward MR analysis (Supplementary Tables 18-19), which was performed without requiring observational significance, similar patterns were observed for the overall agreement ratio (55%; 1052 of 1925 pairs) and risk factors (54%; 856 of 1575 pairs). However, the agreement ratio for diseases decreased to 56% (196 of 350 pairs). The highest agreement ratio was observed for MI, 67% (14 of 21 pairs), though this represented a decline compared with the primary analysis. Agreement for prevalent CAD also declined - falling from 88% to 52% - although incident CAD remained stable (67% and 63% in the primary and the secondary analysis, respectively). Across both analyses, FI, PP, AF, HbA1c, eGFR, and SBP consistently showed low agreement (∼50% or lower) (Supplementary Tables 17,19).

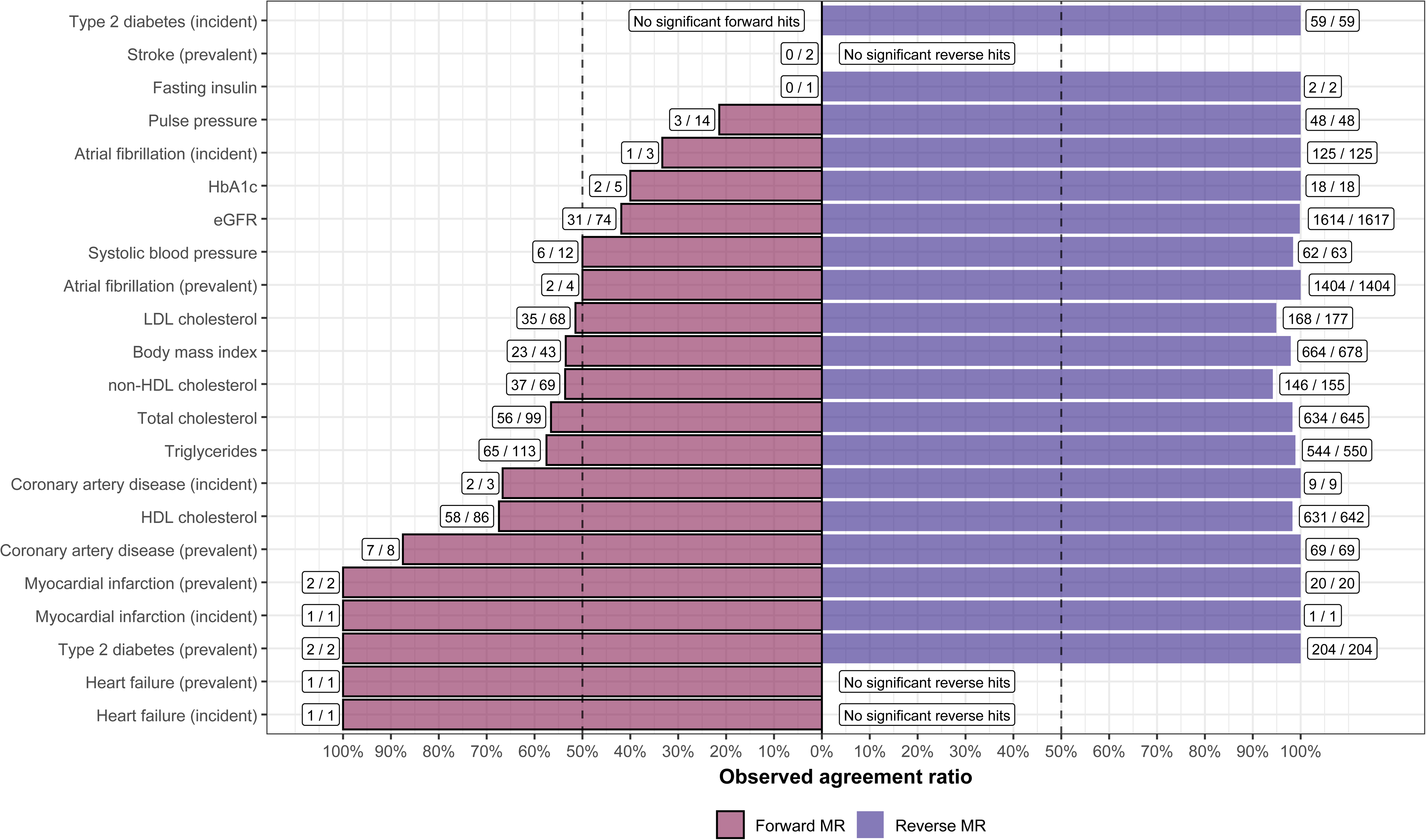

Contrasting to the agreement ratio patterns observed for the forward MR, in the primary reverse MR analysis, 6,422 (99%) of the significant SOMAmer–phenotype pairs showed directional agreement with their observational estimates (Fig. 4A; Supplementary Table 9). Complete agreement was observed across all 1,891 pairs associated with diseases. Among risk factors, mean agreement was 99% (4,531 of 4,595 pairs), ranging from 94% (non-HDL cholesterol) to 100% (FI, HbA1c, PP). The secondary reverse MR estimates were similarly in strong agreement with observational estimates, with 94% (7210 of 7708 pairs) being directionally concordant (Supplementary Table 12).

### Forward MR instrument characteristics and their relationship with observed directional agreement

We evaluated two metrics to assess whether forward MR instrument characteristics influenced directional agreement: coding-variant burden and instrument strength, measured by the harmonic mean F-statistic (HMF). Among 611 SOMAmer–phenotype pairs identified in the primary forward MR analysis, 204 (33%) had instruments that included at least one coding variant (Supplementary Tables 15-16). Coding variants showed a strong association (*χ*^2^test P: 1.9 x 10^-6^) with directional agreement: 69% (140 of 204) of pairs with coding variants were directionally concordant compared with 48% (195 of 407) of pairs without them. This pattern persisted across coding-variant burdens (Supplementary Table 17,), although only a minority of pairs (15%; 30 of 204) carried three or more coding variants.

The distribution of HMF values (median: 33, IQR: 28-40) was comparable across risk factors (median: 33, IQR: 28-40) and diseases (median: 33, IQR 30-38) (Supplementary Table X). Agreement ratios stratified by HMF deciles (Supplementary Table X) did not exhibit any consistent trend, indicating no clear relationship between instrument strength and directional agreement.

These patterns were replicated in the secondary forward MR analysis (Supplementary Table 19). Again, 33% (632 of 1,950) pairs had at least one coding variant, and pairs with coding variants showed higher agreement (63%; 397 of 632) than pairs without coding variants (51%; 655 of 1,293), with a significant *χ*^2^test (P = 3.0 x 10^-6^). As in the primary analysis, agreement varied across HMF deciles but showed no systematic pattern.

Finally, we explored the relationship between coding variant instruments and directional agreement across different strata of SOMAmer-phenotype pairs (Supplementary Table 20). Agreement was consistently higher in the presence of coding variants at both the phenotype group (Figure 5A) and phenotype levels (Figure 5B). For example, agreement for TC increased from 44.4% for pairs without coding variant instruments to 71.1% for those with, and likewise from 51.0% to 71.0% in the lipid group. Similarly, agreement was higher across nearly all deciles of the HMF when coding variants were present (Figure 5C).

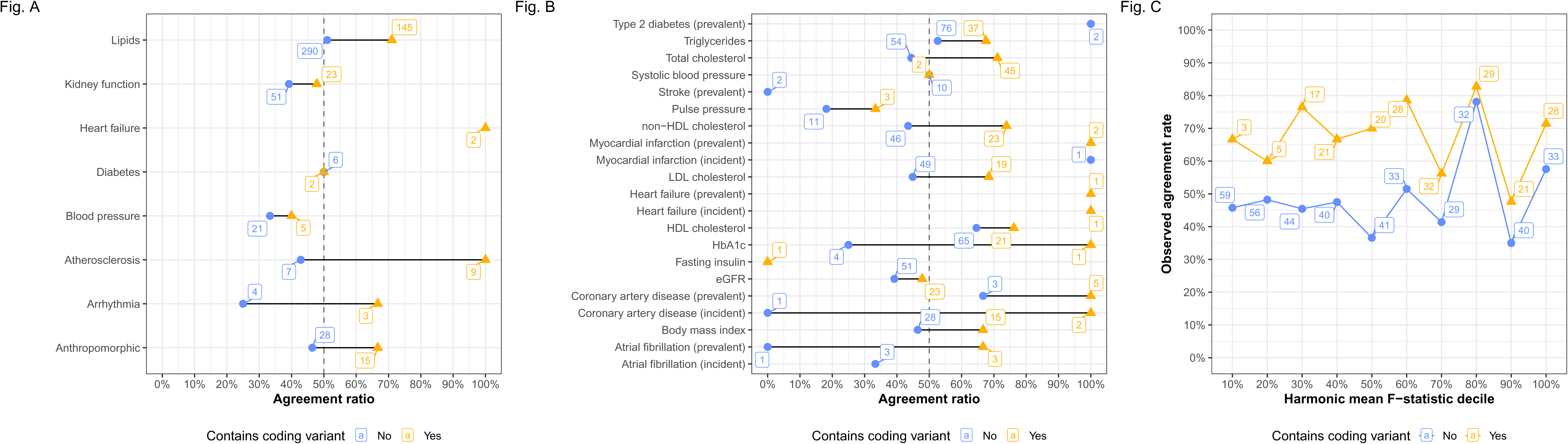

### Posterior estimates of directional agreement between observational and forward MR analyses

To quantify the effect of instrument characteristics on directional agreement and formally evaluate the consistency between observational and MR estimates, we used a hierarchical Bayesian model (Supplementary Methods). Coding variant burden was the only model covariate that influenced directional agreement (Supplementary Figures 3-4), with a posterior median of 0.94 (95% CI: 0.30-1.58) on the log-odds scale (Figure 6A), indicating that greater coding variant burden increases the probability of directional agreement.

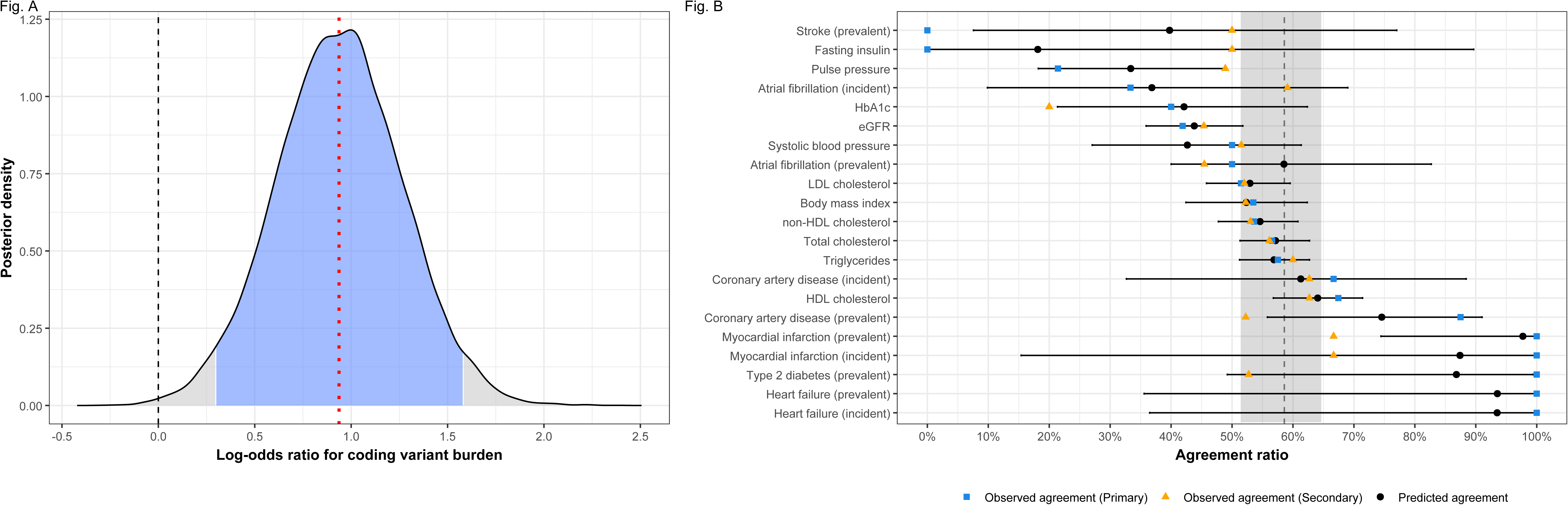

Posterior distributions of phenotype-level directional agreement were estimated from the 611 SOMAmer-phenotype pairs from the primary forward MR analysis (Fig. 6B; Supplementary Table 21). Overall, the global region, defined by the distribution of phenotype-level posterior medians, ranged from 51% to 65% (median 59%) (Fig. 6B), with five observed agreement ratios falling within this range. However, 19 of 21 phenotype-level credible intervals (CIs) intersected the global region, indicating that extremely low or high agreement is mainly driven by sample size, as also indicated by the span of CIs. This interpretation was supported by the comparison with the secondary MR ratios, which were based on a larger number of causal candidates per phenotype. Here, the observed agreement was closer to the expected agreement as predicted by the model for almost all phenotypes (Fig. 6B, Supplementary Table 21). For example, the observed agreement for incident AF increased from 33% in the primary MR to 59% in the secondary MR, when the number of pairs was increased from 3 to 22. Thus, the model captures the tendency of agreement ratios to converge to 59% as sample size increases, reflecting the expected agreement of observational and forward MR estimates for cardiometabolic traits.

## Discussion

Here we analyzed proteomic signatures across 25 cardiometabolic phenotypes in the AGES-RS cohort using 7,288 proteomic measurements, comparing the overlaps and directions of effect for observational and MR associations. To our knowledge, this is the first study to systematically model and quantify such directional agreement across multiple cardiometabolic traits. We conducted two types of MR analyses: a primary analysis restricted to observationally significant proteins, and a secondary analysis that included all proteins with available cis-pQTLs, regardless of observational significance. We observed that generally, observational significance did not enrich for causal candidacy – but rather reverse causation – and across both analyses, agreement between observational and MR estimates was consistently limited in the forward direction but uniformly high in the reverse direction. Coding variant burden emerged as a key factor influencing improved agreement in the forward MR analysis.

In the primary forward MR, cardiometabolic diseases appeared to show higher rates of directional agreement (median 74%) between observational and MR estimates than risk factors (median 54%). However, our model demonstrated that these apparent differences were often inflated by small sample sizes (i.e., few causal candidates). For instance, prevalent CAD showed an observed agreement ratio of 88% (7 out of 8 SOMAmer associations), initially suggesting strong concordance between observational and MR estimates. However, the model’s posterior median agreement was notably lower at 75%, with a wide 95% CI of [56%, 91%], reflecting shrinkage toward the estimated global agreement level (median: 59%; 95% CI: [51%, 65%]). This shrinkage effect, common across phenotypes, underscores how high agreement rates based on sparse signals may overstate true concordance. Importantly, the model’s global region also aligned well with observed agreement ratios derived from the secondary analysis, suggesting that the expected directional agreement between observational and forward MR estimates is approximately 59% - above random chance, but not suggestive of parallel causal mechanisms.

In contrast to the weak alignment observed in the forward direction, reverse MR estimates were nearly always concordant with their corresponding observational associations. This convergence of the two types of signals across multiple traits suggests that circulating protein signatures are strongly shaped by reverse causation. That is, rather than reflecting upstream causal effects of protein levels on disease, the proteomic signal appears to capture downstream biological responses to disease processes. Although our use of incident disease cases helps identifying early proteomic shifts by ensuring that protein measurements precede clinical diagnosis, it remains difficult to disentangle causal perturbations from early prodromal alterations in the proteome. Longitudinal studies with repeated proteomic profiling, particularly in younger and pre-symptomatic individuals, will be essential for resolving the temporal dynamics of these protein changes and clarifying their relevance to disease development^19^. For phenotypes without significant reverse MR associations, it remains unclear whether this absence reflects limited power in the corresponding GWAS or a true lack of reverse causation.

Coding variants were present among cis-instruments for one-third of proteins and their inclusion was consistently associated with higher directional agreement rates. When we stratified by phenotype, phenotype group, and instrument strength, the coding variant strata showed markedly better agreement than their non-coding counterparts. This pattern was reinforced by our Bayesian model, which identified coding variant burden as a strong determinant of increased directional agreement between observational and forward MR estimates. A likely explanation is that coding variants directly affect protein structure and function, yielding more biologically relevant genetic proxies for circulating protein levels^2^. They also provide critical mechanistic context that supports interpretation and triangulation of causal pathways, even when statistical signals are ambiguous. A clear example is PCSK9, whose SNP-profile included rs11591147 (R46L), a well-characterized loss-of-function variant strongly associated with lower LDL cholesterol and reduced coronary heart disease risk^20^. This variant played a central role in guiding the development of PCSK9 inhibitors^21^, illustrating how functional coding variants can clarify biological mechanisms under directional uncertainty.

While these findings underscore the value of coding variants, they also draw attention to broader challenges in interpreting cis-pQTLs within the MR framework. Protein levels in the bloodstream are shaped by a dynamic system influenced by factors such as upstream production, including tissue-specific expression, secretion, leakage and clearance, as well as downstream responses to disease. Approximately 40% of cis-pQTLs also act as expression QTLs across various tissues^2^, and many likely influence protein expression beyond circulating levels^22^, raising concerns about molecular pleiotropy and potential bias in MR estimates^23^. Furthermore, missense cis-pQTLs may affect the epitope recognized by affinity reagents such as SOMAmers, resulting in spurious associations that reflect altered binding affinity rather than true differences in protein concentration^4,24^. A recent structural analysis estimated that ∼12% of cis-pQTLs may be affected by such epitope effects^25^. However, if and how these measurement artifacts can influence directional agreement between observational and MR estimates remains unclear. Together, these considerations suggest that while cis-pQTL instruments may help identify putative causal candidates for complex traits, directional effects observed in plasma or serum may not reliably capture protein activity in disease-relevant tissues where causal mechanisms are likely to operate.

Of the 1,925 significant SOMAmer-phenotype pairs identified in the secondary forward MR analysis that was independent of observational significance, 1,312 (68%) were added compared to the primary MR analysis, while 595 (97%) of the 611 pairs from the primary analysis were recovered. This high degree of overlap likely reflects the enrichment of cis-pQTL instruments among observationally significant SOMAmers. In other words, observational filtering did not isolate a distinct set of causal candidates, as could be expected when correcting for a smaller burden of multiple testing, but rather redundantly identified proteins that would have been tested anyway. Furthermore, bidirectional associations were disproportionately concentrated among observationally prioritized proteins (31% vs. 10%). If the prevalence of bidirectional associations is taken as a measure of causal ambiguity or reverse causation, then this pattern suggests that observational filtering does not preferentially identify proteins with clear, directional effects. Instead, observational associations seem to be more likely to capture proteins influenced by disease processes or pleiotropy, complicating causal interpretation.

Overall, our findings indicate that prioritizing observationally significant proteins for causal analysis (a strategy we ourselves have previously applied) may offer limited benefit. This approach substantially narrows the proteomic search space to a subset that is both redundant and enriched for ambiguous bidirectional associations and reverse causation. Moreover, the apparent improvement in directional agreement exhibited by select phenotypes was largely attributable to sampling variance arising from sparse signals, rather than genuine alignment between observational and genetic effects. Given these limitations, performing MR without prior observational filtering, even when individual-level data is available, may be a more prudent strategy for discovery-driven analyses, despite representing a departure from the method’s original rationale^8^. This approach has become increasingly common in studies of molecular traits such as proteomics^11^, DNA methylation^26^, and metabolomics^27^, whether out of necessity due to the absence of matched observational data, or as a deliberate choice to broaden the search for causal effects. However, omitting this initial filtering does not absolve researchers of the responsibility to ensure that core assumptions of MR are met to the extent that is possible, that they remain aware of the methods limitations, and that they understand how alternative approaches can complement MR results. For example, MR is vulnerable to false positives, which can arise even when no genetic variant is strongly associated with the outcome, whereas colocalization is generally more robust to this issue^7^.

Our study suggests several important directions for future research. One is to examine whether the agreement patterns we observed extend to other populations and phenotypes beyond cardiometabolic traits. Findings from the reverse MR analysis suggest that longitudinal proteomic profiling, particularly in younger and pre-symptomatic populations, may be critical for distinguishing early causal effects from disease-driven changes in protein levels^19^. Improved tissue-specific mapping of pQTLs is also needed to clarify whether MR estimates reflect circulating biology or localized activity in disease-relevant tissues. Another key area for investigation is the role of non-coding variants in proteomic MR, which showed lower directional agreement in our analysis, potentially due to regulatory complexity or pleiotropy. This line of inquiry can be extended to the inclusion of trans-acting variants to the analysis, which we did not consider in this study, and may help uncover previously unrecognized mechanisms of action, including complex network-level effects on disease^28,29^, but at the same time may be more vulnerable to pleiotropic effects. Finally, integrating proteomic MR with complementary omics layers such as transcriptomics, metabolomics, and epigenomics, will help determine whether the patterns of agreement observed here are unique to proteins or reflect broader principles of molecular causality in cardiometabolic disease.

In conclusion, utilizing cis-pQTLs for circulating proteins provides a practical foundation for proteomic MR and the identification of putative causal candidates. However, the limited overlap and weak alignment between observational and forward MR estimates suggests these approaches reflect orthogonal pathways. The strong influence of reverse causation, along with the widespread pleiotropic activity of cis-pQTLs, cautions against interpreting serum protein levels as direct causal exposures. In contrast, the presence of coding variants may offer higher resolution into causal mechanisms, due to their direct effects on protein structure and function. Finally, restricting causal analysis to observationally significant proteins proved limiting and redundant, disproportionately enriching for ambiguous causal associations.

## Methods

### Study population

AGES-RS is a population-based cohort study comprising 5,764 elderly Icelanders, conducted between 2002 and 2006^16^. The participants, aged 66 to 96 years (mean 76 years), were recruited from the Reykjavik Study, which was initiated decades earlier^16^. As part of AGES-RS, participants underwent extensive phenotyping, including questionnaires, physiological measurements, imaging studies, and laboratory assessments, over a three-day period; further details of the study design have been published previously^16^. The study was approved by the Icelandic National Bioethics Committee (VSN-00-063) in accordance with the Helsinki Declaration and the Institutional Review Board of the Intramural Program of the National Institute for Aging. All participants provided informed consent before study enrollment.

### Cardiometabolic phenotypes

We categorized 25 cardiometabolic traits into two groups: prevalent and incident cardiometabolic diseases (dichotomous phenotypes) and risk factors (continuous phenotypes) (Supplementary Table 1). The cardiometabolic diseases included atrial fibrillation (AF), coronary artery disease (CAD), heart failure (HF), stroke, and type 2 diabetes (T2D). The risk factors comprised body mass index (BMI), systolic blood pressure (SBP), diastolic blood pressure (DBP), pulse pressure (PP), high-density lipoprotein (HDL) cholesterol, low-density lipoprotein (LDL) cholesterol, non-HDL cholesterol, total cholesterol (TC), triglycerides (TG), fasting glucose (FG), fasting insulin (FI), HbA1c, and estimated glomerular filtration rate (eGFR) (Supplementary Table 2).

Disease prevalence was assessed at each participant’s entry into AGES-RS. Similarly, follow-up for incident diseases began at study entry and continued for five years for incident T2D, eight years for incident HF, and twelve years for incident AF, CHD, MI, and stroke. Detailed definitions of disease status (prevalent and incident) and biomarker quantification have been described in greater detail in previous publications^2,15,16,30–32^.

For modeling purposes, each phenotype was categorized into one of eight groups: Arrhythmia (prevalent and incident AF), Atherosclerosis (prevalent and incident CHD, MI, and stroke), Heart failure (prevalent and incident HF), Diabetes (FG, FI, HbA1c, and prevalent and incident T2D), Anthropometric (BMI), Blood pressure (PP, DBP, SBP), Lipids (LDL, HDL, non-HDL, TC, TG), and Kidney function (eGFR) (Supplementary Table 2).

### Quantification of serum proteins and genetic data

Serum levels of 6,399 human proteins, targeted by 7,288 slow off-rate modified aptamers (SOMAmers)^33^, were measured using the SomaScan 7k (v4.1)^17^ platform at SomaLogic Inc. (Boulder, US) in 5,364 samples from the AGES-Reykjavik baseline visit. Sample collection and processing for protein measurements were randomized, and all samples were run as a single set.

The SOMAmers had a mean and median intra- and inter-assay coefficient of variation (CV) < 4%. Systematic variability in detection was corrected using hybridization controls and calibrator samples at three dilution levels (20%, 0.5%, and 0.005%), ensuring accurate quantification of protein concentrations via fluorescence intensity. Adaptive normalization by maximum likelihood was performed at SomaLogic Inc. Serum protein measurements were transformed using the Box-Cox transformation and normalized to a mean of 0 and a standard deviation of 1. The 99th percentile measurement value across proteins was identified, and the 99.5th percentile was set as the extreme outlier criterion across all proteins. This resulted in the removal of protein measurements of approximately 4 standard deviations from the mean (absolute value 4.37), affecting an average of 11 samples per protein. Consistent target specificity of the aptamers has been demonstrated through direct validation using mass spectrometry as well as indirect supporting evidence^2^.

The AGES-RS cohort was genotyped using the Illumina hu370CNV array and the Illumina Infinium Global Screening Array. Variants with a call rate <95% or Hardy-Weinberg equilibrium (HWE) P-value <1×10⁻□ were removed prior to imputation. Both arrays were imputed to the Trans-Omics for Precision Medicine (TOPMed r2) panel. After quality control (R^2^<0.7, minor allele frequency < 0.01, HWE P <1×10⁻□), 11,567,385 single nucleotide polymorphisms (SNPs) for 5,368 individuals were available for analysis. The Single Nucleotide Polymorphism Database (dbSNP) version 156^34,35^ was used to classify SNPs as coding (non-synonymous frameshift, non-synonymous missense, non-synonymous nonsense, and synonymous) or noncoding.

### Identification of observational cardiometabolic proteomic signatures

Regression analyses were performed to test for significant associations between SOMAmers and cardiometabolic phenotypes (Figure 1). Multivariable linear regression was employed for risk factors and multivariable logistic regression and Cox proportional hazards regression for prevalent and incident cardiometabolic disease, respectively. All models were adjusted for age, sex, and eGFR levels, except when eGFR was the outcome, in which case models were adjusted for age and sex only. Measured DBP and SBP of participants on antihypertensive medication were adjusted by adding 10 mmHG and 15 mmHG to each respective measurement^3^. The use of statins was accounted for when total, LDL, HDL, and nonHDL cholesterol were regressed on serum protein levels^3^. SOMAmers reaching phenotypic significance (P < 0.05/7,288) were considered part of the phenotype’s proteomic signature.

Finally, Fisher’s exact tests and logistic regression models were fitted across phenotypes to estimate (1) the effect of cis-instrument presence on the probability of observational significance and (2) the effect of observational significance on MR significance.

### Causal inference by means of bidirectional, two-sample Mendelian randomization

A bidirectional, two-sample Mendelian randomization (MR) analysis^8^ was conducted to evaluate the causal candidacy of serum proteins across cardiometabolic phenotypes (Figure 1). The MR analyses were performed in two phases. The primary analysis focused on SOMAmer–phenotype pairs that demonstrated significant observational associations, with forward MR additionally requiring the presence of cis-acting instruments (defined below). The secondary analysis applied the same MR framework without filtering for observational significance. Summary statistics for genetic liability to 25 cardiometabolic phenotypes were obtained from 11 external GWAS^36–46^ (Supplementary Table 6).

In the forward MR analysis (exposure: serum protein levels; outcome: cardiometabolic phenotype), protein quantitative trait loci (pQTL) SNPs located within a cis-region spanning ±500 kb of the protein-coding gene (targeted by its respective SOMAmer) were selected as potential instruments. SNPs were clumped for linkage disequilibrium (LD) using PLINK v1.9^47^ with a threshold of r² ≥ 0.2 (window ±500 kb), allowing for inclusive SNP selection within these constrained regions^5^. Instruments were further filtered based on cis-window significance (P < 0.05 / number of SNPs before clumping) and instrument strength (F-statistic > 10). Correlation between retained SNPs was accounted for in the MR analysis using LD-adjusted methods^5,48^ (see below). Where protein-associated SNPs were absent in external GWAS datasets, proxy variants (r² > 0.8) were used when available. In the reverse MR analysis (exposure: cardiometabolic phenotype; outcome: serum protein levels), genome-wide significant SNPs (P < 5 × 10⁻□) were selected for each phenotype and clumped (LD-threshold: r² > 0.01, window ±10,000 kb). To mitigate bias from cis-mediated protein expression, SNPs located within the cis-region (±500 kb of the corresponding protein-coding gene) were excluded from the reverse analysis. All SNP sets were harmonized using the TwoSampleMR R package^49^.

Single-SNP and multi-SNP causal estimates were computed using Wald’s ratio estimator and the generalized weighted least squares (GWLS) estimator, respectively. The GWLS estimator was applied in both forward and reverse MR settings as it accounts for LD structure in the cis-region (forward MR) and simplifies to the inverse variance-weighted (IVW) estimator when SNPs are uncorrelated (reverse MR)^5,50^. MR estimates reaching phenotype-level significance (Benjamini-Hochberg FDR < 0.05) were retained for further analysis. Instrument strength was quantified using the harmonic mean of the F-statistics (HMF) for the SNPs included in each MR estimate. Similarly, the total coding variant burden (TC) was defined as the number of coding variants among the SNPs comprising each genetic instrument.

### Assessment of instrument heterogeneity

To evaluate the robustness of causal effect estimates from the primary forward MR analysis, we conducted a sensitivity analysis assessing SNP-level heterogeneity within cis-instrument sets (Figure 1). For SOMAmer–phenotype pairs with more than one SNP instrument, we applied a Bayesian random-effects model, treating SNP-specific Wald ratio estimates as arising from a shared causal effect θ with additional between-SNP variability r. The model incorporated the LD-adjusted covariance matrix of the Wald ratios to account for correlation between SNPs (Supplementary Methods). Causal estimates that remained directionally consistent and whose 95% credible intervals (CI) excluded zero were considered to have passed the heterogeneity test. The model was implemented in Stan using the rstan R package^51^. We ran four Markov chain Monte Carlo (MCMC) chains with 5,000 warm-up and 5,000 post-warm-up iterations per chain. Convergence was assessed using the potential scale reduction factor *Ȓ*.

### Modeling of agreement ratios

We modeled directional agreement between observational and forward MR estimates using a hierarchical Bayesian logistic regression model (Supplementary Figure 1). The model estimated the probability of directional agreement for each SOMAmer–phenotype pair while accounting for phenotype group, phenotype-specific effects, protein-level effects, SOMAmer correlations, instrument strength, and coding variant burden (Supplementary Methods). The model was implemented in Stan with the rstan R package^51^ using four MCMC chains of 5,000 warm-up and 5,000 post-warm-up iterations each. Convergence was assessed using the potential scale reduction factor *Ȓ*. Posterior summaries were based on 10,000 draws. Phenotype-level agreement estimates were computed by marginalizing over posterior agreement probabilities for each phenotype. Observed agreement ratios from the secondary MR analysis were not used to fit the model but were compared post hoc to the predicted intervals and global region.

## Data Availability

Data from the AGES-Reykjavik study are available through collaboration under a data usage agreement with the IHA. All access to data is controlled via the use of a subject-signed informed consent authorization. All other data supporting the conclusions of the paper are presented in the main text and freely available as a supplement to this manuscript.

## References

1. Sun, B. B. et al. Genomic atlas of the human plasma proteome. Nature 558, 73–79 (2018).

2. Emilsson, V. et al. Co-regulatory networks of human serum proteins link genetics to disease. Science 361, 769–773 (2018).

3. Gudjonsson, A. et al. A genome-wide association study of serum proteins reveals shared loci with common diseases. Nat. Commun. 13, 480 (2022).

4. Suhre, K., McCarthy, M. I. & Schwenk, J. M. Genetics meets proteomics: perspectives for large population-based studies. Nat. Rev. Genet. 22, 19–37 (2021).

5. Gkatzionis, A., Burgess, S. & Newcombe, P. J. Statistical methods for cis-Mendelian randomization with two-sample summary-level data. Genet. Epidemiol. 47, 3–25 (2023).

6. Gill, D. et al. Mendelian randomization for studying the effects of perturbing drug targets. Wellcome Open Res. 6, 16 (2021).

7. Zuber, V. et al. Combining evidence from Mendelian randomization and colocalization: Review and comparison of approaches. Am. J. Hum. Genet. 109, 767–782 (2022).

8. Davey Smith, G. & Ebrahim, S. ‘Mendelian randomization’: can genetic epidemiology contribute to understanding environmental determinants of disease?*. Int. J. Epidemiol. 32, 1–22 (2003).

9. Davey Smith, G. & Hemani, G. Mendelian randomization: genetic anchors for causal inference in epidemiological studies. Hum. Mol. Genet. 23, R89–R98 (2014).

10. Chen, L. et al. Systematic Mendelian randomization using the human plasma proteome to discover potential therapeutic targets for stroke. Nat. Commun. 13, 6143 (2022).

11. Zheng, J. et al. Phenome-wide Mendelian randomization mapping the influence of the plasma proteome on complex diseases. Nat. Genet. 52, 1122–1131 (2020).

12. Zhang, K. et al. Proteome-Wide Mendelian Randomization Identifies Therapeutic Targets for Abdominal Aortic Aneurysm. J. Am. Heart Assoc. 14, e038193 (2025).

13. Titova, O. E. et al. Plasma proteome and incident myocardial infarction: sex-specific differences. Eur. Heart J. 45, 4647–4657 (2024).

14. Schuermans, A. et al. Integrative proteomic analyses across common cardiac diseases yield mechanistic insights and enhanced prediction. *Nat*. Cardiovasc. Res. 3, 1516–1530 (2024).

15. Gudmundsdottir, V. et al. Circulating Protein Signatures and Causal Candidates for Type 2 Diabetes. Diabetes 69, 1843–1853 (2020).

16. Harris, T. B. et al. Age, Gene/Environment Susceptibility – Reykjavik Study: Multidisciplinary Applied Phenomics. Am. J. Epidemiol. 165, 1076–1087 (2007).

17. Candia, J., Daya, G. N., Tanaka, T., Ferrucci, L. & Walker, K. A. Assessment of variability in the plasma 7k SomaScan proteomics assay. Sci. Rep. 12, 17147 (2022).

18. Burgess, S., Davies, N. M. & Thompson, S. G. Bias due to participant overlap in two-sample Mendelian randomization. Genet. Epidemiol. 40, 597–608 (2016).

19. Tang, J. et al. Longitudinal serum proteome mapping reveals biomarkers for healthy ageing and related cardiometabolic diseases. Nat. Metab. 7, 166–181 (2025).

20. Cohen, J. C., Boerwinkle, E., Mosley, T. H. & Hobbs, H. H. Sequence variations in PCSK9, low LDL, and protection against coronary heart disease. N. Engl. J. Med. 354, 1264–1272 (2006).

21. Sabatine, M. S. et al. Evolocumab and Clinical Outcomes in Patients with Cardiovascular Disease. N. Engl. J. Med. 376, 1713–1722 (2017).

22. Fang, H. et al. Regulation of protein abundance in normal human tissues. MedRxiv Prepr. Serv. Health Sci. 2025.01.10.25320181 (2025) doi:10.1101/2025.01.10.25320181.

23. Hemani, G., Bowden, J. & Davey Smith, G. Evaluating the potential role of pleiotropy in Mendelian randomization studies. Hum. Mol. Genet. 27, R195–R208 (2018).

24. Suhre, K. et al. A genome-wide association study of mass spectrometry proteomics using a nanoparticle enrichment platform. Nat. Genet. 1–10 (2025) doi:10.1038/s41588-025-02413-w.

25. Kuliesius, J. et al. Epitope Effect Prevalence in Affinity-based pQTL studies. 2025.06.20.660695 Preprint at 10.1101/2025.06.20.660695 (2025).

26. Schuurmans, I. K., Dunn, E. C. & Lussier, A. A. DNA methylation as a possible mechanism linking childhood adversity and health: results from a 2-sample mendelian randomization study. Am. J. Epidemiol. 193, 1541–1552 (2024).

27. Wong, T. H. T. et al. A two-sample Mendelian randomization study explores metabolic profiling of different glycemic traits. *Commun*. Biol. 7, 293 (2024).

28. Liu, X., Li, Y. I. & Pritchard, J. K. Trans Effects on Gene Expression Can Drive Omnigenic Inheritance. Cell 177, 1022–1034.e6 (2019).

29. Boyle, E. A., Li, Y. I. & Pritchard, J. K. An Expanded View of Complex Traits: From Polygenic to Omnigenic. Cell 169, 1177–1186 (2017).

30. Emilsson, V. et al. Proteomic prediction of incident heart failure and its main subtypes. Eur. J. Heart Fail. 26, 87–102 (2024).

31. Jonmundsson, T., et al. A proteomic analysis of atrial fibrillation in a prospective longitudinal cohort (AGES-Reykjavik study). EP Eur. 25, euad320 (2023).

32. Bankier, S. et al. Circulating causal protein networks linked to future risk of myocardial infarction. 2025.02.07.25321789 Preprint at 10.1101/2025.02.07.25321789 (2025).

33. Gold, L. et al. Aptamer-Based Multiplexed Proteomic Technology for Biomarker Discovery. PLOS ONE 5, e15004 (2010).

34. Phan, L. et al. The evolution of dbSNP: 25 years of impact in genomic research. Nucleic Acids Res. 53, D925–D931 (2025).

35. Sherry, S. T. et al. dbSNP: the NCBI database of genetic variation. Nucleic Acids Res. 29, 308–311 (2001).

36. Aragam, K. G. et al. Discovery and systematic characterization of risk variants and genes for coronary artery disease in over a million participants. Nat. Genet. 54, 1803–1815 (2022).

37. Chen, J. et al. The trans-ancestral genomic architecture of glycemic traits. Nat. Genet. 53, 840–860 (2021).

38. Evangelou, E. et al. Genetic analysis of over 1 million people identifies 535 new loci associated with blood pressure traits. Nat. Genet. 50, 1412–1425 (2018).

39. Graham, S. E. et al. The power of genetic diversity in genome-wide association studies of lipids. Nature 600, 675–679 (2021).

40. Hartiala, J. A. et al. Genome-wide analysis identifies novel susceptibility loci for myocardial infarction. Eur. Heart J. 42, 919–933 (2021).

41. Mahajan, A. et al. Multi-ancestry genetic study of type 2 diabetes highlights the power of diverse populations for discovery and translation. Nat. Genet. 54, 560–572 (2022).

42. Mishra, A. et al. Stroke genetics informs drug discovery and risk prediction across ancestries. Nature 611, 115–123 (2022).

43. Nielsen, J. B. et al. Biobank-driven genomic discovery yields new insight into atrial fibrillation biology. Nat. Genet. 50, 1234–1239 (2018).

44. Pulit, S. L. et al. Meta-analysis of genome-wide association studies for body fat distribution in 694 649 individuals of European ancestry. Hum. Mol. Genet. 28, 166–174 (2019).

45. Shah, S. et al. Genome-wide association and Mendelian randomisation analysis provide insights into the pathogenesis of heart failure. Nat. Commun. 11, 163 (2020).

46. Stanzick, K. J. et al. Discovery and prioritization of variants and genes for kidney function in >1.2 million individuals. Nat. Commun. 12, 4350 (2021).

47. Purcell, S. et al. PLINK: a tool set for whole-genome association and population-based linkage analyses. Am. J. Hum. Genet. 81, 559–575 (2007).

48. Emilsson, V. et al. A proteogenomic signature of age-related macular degeneration in blood. Nat. Commun. 13, 3401 (2022).

49. Hemani, G. et al. The MR-Base platform supports systematic causal inference across the human phenome. eLife 7, e34408 (2018).

50. Burgess, S., Dudbridge, F. & Thompson, S. G. Combining information on multiple instrumental variables in Mendelian randomization: comparison of allele score and summarized data methods. Stat. Med. 35, 1880–1906 (2016).

51. Stan Development Team. RStan: the R interface to Stan.

