## Supplementary material for "Systematic comparison of observational and Mendelian Randomization estimates for cardiometabolic proteomic signatures": Figure Captions

### Figure legends

**Figure 1: Overview of analysis workflow.** The observed proteomic signatures of 25 cardiometabolic phenotypes were identified using regression models. Bidirectional, two-sample MR was used to evaluate potentially causal associations: a primary analysis restricted to observationally significant SOMAmer-phenotype pairs, and a secondary analysis including all pairs. Directional agreement between observational and MR estimates was quantified for each phenotype and modeled hierarchically to capture global and phenotype-specific patterns.

**Figure 2: Proteomic signatures of cardiometabolic phenotypes in AGES-RS.** Phenotypes are grouped as either diseases (blue fill, black border) or risk factors (yellow fill, no border). **(A)** Number of SOMAmer-phenotype pairs reaching observational significance (P < 0.05/7,288) for each phenotype, representing the proteomic signature for each trait. White box labels indicate the counts underlying each bar. **(B)** Proportion of observationally significant SOMAmers that had at least one cis-instrument available. The broken line denotes the overall proportion of SOMAmers with cis-acting instruments in AGES-RS (28%; 2,062/7,288). White box labels indicate the counts underlying each proportion.  **(C)** Forest plot showing enrichment of SOMAmers with cis-instruments within the proteomic signatures. Each crossbar shows the Wald 95% confidence interval for the log-odds ratio (solid black circle if P < 0.05, open circle otherwise) of cis-instrument presence. Point estimates greater than 0 indicate enrichment of cis-SOMAmers, and those less than 0 indicate depletion.

**Figure 3: Causal profiling of proteomic signatures in AGES-RS. (A)** Bar plot illustrating the proportion of significant (FDR < 0.05) SOMAmer-phenotype pairs from the primary MR analysis, where pairs were pre-filtered for observational significance. Proportions from the forward analysis (SOMAmer → phenotype) are shown on the left (pink fill, black border) and those from the reverse analysis (phenotype → SOMAmer) on the right (purple fill, no border). White box labels represent the counts underlying each proportion. **(B)** The same as (A), but for the secondary MR analysis, where all SOMAmer-phenotype pairs were considered. **(C)** Forest plot showing the effect of observational significance on forward-MR significance. For each phenotype (disease: blue filled, black border; risk factor: yellow-filled, no border), the crossbar shows the Wald 95% confidence interval for the log-odds ratio (solid black if P < 0.05, open circle otherwise) of forward-MR significance given observational significance. White box labels display FET P-values for phenotypes unable to be fitted with logistic regression. **(D)** The same as (C), but for reverse-MR significance. White box labels display either FET P-values for phenotypes unable to be fitted with logistic regression or indicate if the phenotype could not be tested because no significant reverse-MR estimates were observed.

**Figure 4: Observed and posterior estimates of directional agreement between observational and MR estimates.** Bar plot showing the observed proportion of directional agreement between observational and MR estimates for SOMAmer-phenotype pairs identified in the primary MR analysis, where pairs were pre-filtered for observational significance. Proportions from the forward analysis (SOMAmer → phenotype) are shown on the left (pink fill, black border) and those from the reverse analysis (phenotype → SOMAmer) on the right (purple fill, no border). The broken line at 50% denotes the expected agreement by chance. White box labels show the counts underlying each proportion or state if the phenotype had no significant MR estimates (FDR < 0.05).

**Figure 5: The effect of coding variant burden on directional agreement for forward MR estimates.** **(A)** Observed directional agreement by phenotype group, stratified by coding variant status. **(B)** Observed directional agreement by phenotype, stratified by coding variant status. **(C)** Observed directional agreement of harmonic mean F-statistic (HMF), stratified by coding variant status**.** In the panels, blue circles represent agreement ratios based on SOMAmer-phenotype pairs without coding variants, and yellow triangles represent ratios for pairs with coding variants. Similarly, blue-bordered (without coding variants) and yellow-bordered (with coding variants) boxes represent the sample size underlying each point.

***Figure 6: The effect of coding variant burden on directional agreement for forward MR estimates.*** *(****A)*** *Posterior distribution of the effect of coding variant burden on directional agreement. The shaded blue region indicates the 95% credible interval (CI), and the gray regions represent the posterior mass outside the CI. The red dotted line marks the posterior median (0.94) on the log-odds scale.* ***(B)*** *Forest plot showing the posterior estimates of directional agreement for each of the 21 phenotypes with significant forward-MR estimates (FDR < 0.05) identified in the primary analysis. Black circles represent the posterior median probability of directional agreement, and solid black lines denote the corresponding 95% credibility intervals. Observed agreement ratios from the primary MR analysis are shown as blue squares, and those from the secondary MR analysis (when available) as yellow triangles. The vertical dashed line marks the global posterior median agreement ratio (59%), and the gray band shows it 95% credible interval (51%-65%), or the global region.*
