## Supplementary methods for "Systematic comparison of observational and Mendelian Randomization estimates for cardiometabolic proteomic signatures"

### Supplementary Methods (draft)

##### Statistical analyses

###### Enrichment of cis-SOMAmers in Bonferroni tail

Our goal is to assess whether the presence of a cis-instrument increases the probability that a SOMAmer is observationally significant. For each phenotype $p$, let $i = 1, \ldots, n_p$ index SOMAmers included in the observational analysis. For each SOMAmer $i$, let:

- $P_i$ denote the $p$-value from the observational association test (linear regression, logistic regression, Cox proportional hazard regression).
- $X_i = I(P_i < \alpha_B)$ be an indicator of Bonferroni significance ($\alpha_B = 0.05/7288$).
- $Y_i = I(\text{SOMAmer } i \text{ has } \geq 1 \text{ cis-acting variant})$ be an indicator of cis-instrument status.

The conditional probability:

$$\pi = P(X_i = 1 | Y_i = 1),$$

where enrichment (higher odds of significance among cis-SOMAmers) corresponds to $\pi > P(X_i = 1 | Y_i = 0)$. We use logistic regression to estimate $\pi$. Namely:

$$\text{logit} \frac{\pi}{1-\pi} = \alpha + \beta Y_i,$$

where:

- $\alpha$ is the intercept, and
- $\beta$ is the log-odds ratio comparing cis versus non-cis SOMAmers, and $\beta > 0$ indicates enrichment.

In R:

```
glm(X_i ~ Y_i, data = df, family = binomial())
```

##### Enrichment of MR significance

Our goal is to assess whether observational significance increases the probability of MR significance. The discussion below focuses on forward MR significance, but applies equally to reverse MR significance. For each phenotype $p$, we only consider SOMAmers that have at least one cis-instrument (which are eligible for forward MR). Index these SOMAmers by $i = 1, \dots, n_p$ and for each SOMAmer $i$, let:

- $P_i^{\mathrm{o}}$ denote the observational association P-value,
- $P_i$ and $Q_i$ denote the primary forward MR P-value and its respective FDR estimate,
- $P_i'$ and $Q_i'$ denote the secondary forward MR P-value and its respective FDR estimate.

The Bonferroni indicator for each SOMAmer is as above:

$$X_i = I(P_i^o < \alpha_B)$$

Similarly, we have the primary MR and the secondary MR significance indicators:

$$Z_i = I(Q_i < \alpha_{\mathrm{FDR}}), \quad Z_i' = I(Q_i' < \alpha_{\mathrm{FDR}}),$$

where $\alpha_{\mathrm{FDR}} = 0.05$. Combining $Z_i$ and $Z_i'$ yields the MR significance indicator

$$Y_i = I(Z_i + Z_i' > 0),$$

such that:

- $Y_i = 1$ means that the SOMAmer is MR significant in either analysis.
- $Y_i = 0$ neither analysis yielded MR significance.

Again (and by symbolic overload), the conditional probability

$$\pi = P(Y_i = 1|X_i = 1),$$

where enrichment (higher odds of significance among cis-SOMAmers) corresponds to $\pi > P(Y_i = 1|X_i = 0)$, can be estimated with logistic regression:

$$\text{logit}\,\frac{\pi}{1-\pi} = \alpha + \beta X_i,$$

where:

- $\alpha$ is the intercept, and
- $\beta$ is the change in log-odds of (any) MR significance associated with observational significance, and $\beta > 0$ indicates enrichment.

In R:

```
glm(Y_i ~ X_i, data = df, family = binomial())
```

#### Bayesian model accounting for heterogeneity

To estimate the causal effect of a given exposure on an outcome using multiple genetic instruments, we implemented a Bayesian random-effects model that accounts for both sampling uncertainty and heterogeneity across SNP-specific effect estimates. Specifically, we modeled the vector of Wald ratio estimates $\hat{\beta} \in \mathbb{R}^N$ as multivariate normal distributed centered around a common causal effect $\theta$, with covariance given by the sum of the measurement error covariance matrix and a random-effects variance component:

$$\hat{\beta} \sim \mathcal{N}(\theta \mathbf{1}_N, \Sigma + \tau^2 I_N)$$

where:

- $\Sigma \in \mathbb{R}^{N \times N}$ is the known covariance matrix of the Wald ratios, incorporating linkage disequilibrium (LD) between instruments,
- $\tau^2$ is a non-negative variance parameter capturing residual heterogeneity between SNP-specific estimates not explained by sampling error,
- $I_N$ is the identity matrix of size $N$.

Priors were specified as follows:

- $\theta \sim \mathcal{N}(0, 1)$, representing a weakly informative prior on the overall causal effect,

- $\tau \sim \text{Cauchy}(0, 0.1)$, a standard weakly informative half-Cauchy prior on the standard deviation of the random effects.

Inference was performed using Hamiltonian Monte Carlo (HMC) via Stan. We used the Cholesky decomposition of the total covariance matrix $\Sigma + \tau^2 I_N$ for computational efficiency and numerical stability.

```
data {
  int<lower=1> N;                    // Number of SNPs
  vector[N] beta_hat;                // Wald ratios
  matrix[N, N] Sigma;                // Covariance matrix of Wald ratios
}

parameters {
  real theta;                       // Overall causal effect
  real<lower=0> tau;                // Between-SNP heterogeneity (SD)
}

model {
  matrix[N, N] Sigma_total;
  matrix[N, N] L_Sigma_total;

  // Priors
  theta ~ normal(0, 1);
  tau ~ cauchy(0, 0.1);

  // Total covariance = measurement error + heterogeneity
  Sigma_total = Sigma + diag_matrix(rep_vector(tau^2, N));
  L_Sigma_total = cholesky_decompose(Sigma_total);

  // Multivariate normal likelihood
  beta_hat ~ multi_normal_cholesky(rep_vector(theta, N), L_Sigma_total);
}
```

#### Bayesian Hierarchical Logistic Model for Directional Agreement

We further implemented a Bayesian hierarchical logistic regression model to estimate the probability of directional agreement between observational and Mendelian randomization (MR) estimates across SOMAmer–phenotype pairs. The binary response variable $y_i \in \{0, 1\}$ indicated whether pair $i$ demonstrated concordant directions.

The model included hierarchical effects at multiple levels:

- A global intercept $\Delta$,
- Group-specific effects $\gamma_g$ for phenotype group $g$,
- Phenotype-specific effects $\alpha_p$,
- SOMAmer-level effects $\zeta_k$,
- SOMAmer-specific deviations modeled via a Cholesky-correlated random effect $\beta_s$ based on a known SOMAmer correlation matrix,
- Covariate effects for harmonic mean F-statistics ($\theta_f$) and log-transformed coding variant burden ($\theta_{tc}$).

The linear predictor for each observation $i$ was defined as:

$$\text{logit}(\Pr(y_i = 1)) = \Delta + \gamma_{g[i]} + \alpha_{p[i]} + \beta_{s[i]} + \theta_f \cdot \text{F-stat}_i + \theta_{tc} \cdot \log(1 + \text{coding burden}_i)$$

Priors:

- All group-level raw effects were given standard normal priors and scaled by group-specific standard deviations, with exponential hyperpriors:
  - $\gamma_{\text{raw}} \sim \mathcal{N}(0,1)$ with $\sigma_\gamma \sim \text{Exponential}(1)$
  - $\alpha_{\text{raw}} \sim \mathcal{N}(0,1)$ with $\sigma_\alpha \sim \text{Exponential}(1)$
  - $\zeta_{\text{raw}} \sim \mathcal{N}(0,1)$ with $\sigma_\zeta \sim \text{Exponential}(1)$
- Protein-level deviations were modeled as $\beta_s = \zeta[\text{protein}(s)] + \sigma_\varepsilon \cdot (L z_\beta)$, where $z_\beta \sim \mathcal{N}(0,1)$ and $L$ is the Cholesky decomposition of the SOMAmer correlation matrix.
- Covariate effects: $\theta_f \sim \mathcal{N}(0,1)$ and $\theta_{tc} \sim \mathcal{N}(0,0.5)$.

Posterior inference was again performed using HMC in Stan, and the model included a `generated quantities` block to compute observation-level log-likelihoods for use in model comparison and validation.

This hierarchical structure enabled robust inference while accounting for shared variation at the phenotype group, phenotype, and protein levels, as well as inter-SOMAmer correlation.

```
data {
  int<lower=1> N;                  // number of protein-phenotype pairs
  int<lower=1> P;                  // number of phenotypes
  int<lower=1> S;                  // number of somamers
  int<lower=1> G;                  // number of phenotype groups
  int<lower=1> K;                  // number of unique proteins targeted by SOMAmers

  int<lower=1, upper=P> phenotype[N]; // Phenotype at i-th index
  int<lower=1, upper=S> somamer[N];   // SOMAmer at i-th index
  // Maps
```

```
  int<lower=1, upper=G> group_map[P];     // Map phenotype at i-th index to its group
  int<lower=1, upper=K> protein_map[S]; // maps SOMAmer s to protein k
  vector[N] f_stat;   // harmonic f stat
  vector[N] log_tc;   // log(1 + total_coding_variants)

  matrix[S, S] L;   // Cholesky of correlation matrix between SOMAmers

  int<lower=0, upper=1> y[N];
}

parameters {
  real alpha;                       // global intercept

  // Non-centered parameterizations for the hierarchical effects
  vector[G] gamma_raw;              // raw group effects
  real<lower=0> sigma_gamma;

  vector[P] alpha_raw;              // raw phenotype effects
  real<lower=0> sigma_alpha;

  vector[S] z_beta;                // raw somamer effects
  real<lower=0> sigma_eps;

  vector[K] zeta_raw;             // raw protein effects
  real<lower=0> sigma_zeta;

  real theta_f;   // Harmonic F
  real theta_tc;  // Total coding variants behind phenotype-protein pair
}

transformed parameters {
  vector[G] gamma_g = sigma_gamma * gamma_raw;
  vector[P] alpha_p = sigma_alpha * alpha_raw;
  vector[K] zeta = sigma_zeta * zeta_raw;

  vector[S] beta_mu; // Mean somamer effect from zeta
  vector[S] beta_s;

  for (s in 1:S) {
    beta_mu[s] = zeta[protein_map[s]];
  }
  beta_s = beta_mu + sigma_eps * (L * z_beta); // Transformed protein effects
```

```
}

model {
  // Priors
  alpha ~ normal(0, 1);

  gamma_raw ~ normal(0, 1);
  sigma_gamma ~ exponential(1);

  alpha_raw ~ normal(0, 1);
  sigma_alpha ~ exponential(1);

  zeta_raw ~ normal(0, 1);
  sigma_zeta ~ exponential(1);

  z_beta ~ normal(0, 1);
  sigma_eps ~ exponential(1);

  theta_f ~ normal(0, 1);
  theta_tc ~ normal(0, 0.5);

  // Likelihood: vectorized the linear predictor
  {
    vector[N] linpred;
    for (i in 1:N) {
      int p = phenotype[i];
      int g = group_map[p];
      int s = somamer[i];
      linpred[i] = alpha + gamma_g[g] + alpha_p[p] + beta_s[s] +
      theta_f * f_stat[i] +
      theta_tc * log_tc[i];
    }
    y ~ bernoulli_logit(linpred);
  }
}

generated quantities {
  vector[N] log_lik;

  for (i in 1:N) {
    int p = phenotype[i];
    int g = group_map[p];
```

```
    int s = somamer[i];
    log_lik[i] = bernoulli_logit_lpmf(y[i] |
      alpha + gamma_g[g] +
      alpha_p[p] +
      beta_s[s] +
      theta_f * f_stat[i] +
      theta_tc * log_tc[i]
    );
  }
}
```
