## Supplementary material for "Systematic comparison of observational and Mendelian Randomization estimates for cardiometabolic proteomic signatures": Table 1

Table 1: Baseline characteristics of AGES-RS cohort

| **Characteristic** | **Overall** N = 5,364*^1^* | **Men** N = 2,294*^1^* | **Women** N = 3,070*^1^* | **p-value***^2^* |
| --- | --- | --- | --- | --- |
| **Demographics** | | | | |
| Age | 76 (72-81) | 76 (72-81) | 76 (72-81) | 0.3 |
| **Anthropomorphic** | | | | |
| Height | 166 (160-174) | 175 (171-180) | 161 (157-165) | <0.001 |
| Weight | 74 (66-85) | 82 (73-91) | 69 (62-78) | <0.001 |
| Body mass index [BMI] | 26.7 (24.1-29.5) | 26.6 (24.3-29.0) | 26.8 (23.9-29.9) | 0.2 |
| **Physiological** | | | | |
| Diastolic blood pressure [DBP] (mmHG) | 83 (77-90) | 86 (79-92) | 82 (76-89) | <0.001 |
| Systolic blood pressure [SBP] (mmHG) | 155 (143-170) | 156 (144-171) | 155 (142-169) | 0.021 |
| Pulse pressure [PP] (mmHG) | 67 (56-79) | 65 (55-77) | 68 (56-81) | <0.001 |
| non-HDL cholesterol (mmol/L) | 3.99 (3.24-4.77) | 3.74 (3.00-4.48) | 4.17 (3.44-4.93) | <0.001 |
| LDL cholesterol (mmol/L) | 3.47 (2.74-4.18) | 3.23 (2.51-3.89) | 3.62 (2.94-4.36) | <0.001 |
| HDL cholesterol (mmol/L) | 1.53 (1.26-1.85) | 1.35 (1.14-1.63) | 1.68 (1.41-1.99) | <0.001 |
| Total cholesterol [TC] (mmol/L) | 5.60 (4.80-6.40) | 5.20 (4.40-5.90) | 5.90 (5.20-6.61) | <0.001 |
| Triglycerides [TG] (mmol/L) | 1.04 (0.78-1.43) | 1.01 (0.75-1.40) | 1.07 (0.80-1.46) | <0.001 |
| Fasting glucose [FG] (mmol/L) | 5.50 (5.20-6.00) | 5.60 (5.30-6.10) | 5.40 (5.10-5.90) | <0.001 |
| Fasting insulin [FI] (µU/mL) | 8 (6-12) | 9 (6-13) | 8 (5-12) | <0.001 |
| HbA1c (g/dL) | 0.48 (0.43-0.52) | 0.49 (0.45-0.54) | 0.46 (0.43-0.51) | <0.001 |
| eGFR (mL/min/1.73m2) | 64 (54-75) | 66 (56-76) | 63 (53-75) | <0.001 |
| **Atrial fibrillation [AF]** | | | | |
| Prevalent |  |  |  | <0.001 |
| 0 | 4,813 (91%) | 1,956 (86%) | 2,857 (94%) |  |
| 1 | 498 (9.4%) | 320 (14%) | 178 (5.9%) |  |
| Incident |  |  |  | <0.001 |
| 0 | 4,195 (79%) | 1,709 (75%) | 2,486 (82%) |  |
| 1 | 1,116 (21%) | 567 (25%) | 549 (18%) |  |
| Follow-Up | 9.4 (5.5-10.6) | 8.8 (4.4-10.3) | 9.7 (6.5-10.8) | <0.001 |
| **Coronary artery disease [CAD]** | | | | |
| Prevalent |  |  |  | <0.001 |
| 0 | 4,114 (77%) | 1,512 (66%) | 2,602 (86%) |  |
| 1 | 1,197 (23%) | 764 (34%) | 433 (14%) |  |
| Incident |  |  |  | <0.001 |
| 0 | 3,715 (70%) | 1,348 (59%) | 2,367 (78%) |  |
| 1 | 1,596 (30%) | 928 (41%) | 668 (22%) |  |
| Follow-Up | 9.2 (4.4-10.5) | 7.4 (3.2-10.1) | 9.7 (5.8-10.8) | <0.001 |
| **Myocardial infarction [MI]** | | | | |
| Prevalent |  |  |  | <0.001 |
| 0 | 4,658 (88%) | 1,859 (82%) | 2,799 (92%) |  |
| 1 | 653 (12%) | 417 (18%) | 236 (7.8%) |  |
| Incident |  |  |  | <0.001 |
| 0 | 4,485 (84%) | 1,813 (80%) | 2,672 (88%) |  |
| 1 | 826 (16%) | 463 (20%) | 363 (12%) |  |
| Follow-Up | 9.7 (6.4-10.8) | 9.3 (5.3-10.5) | 9.8 (7.4-10.9) | <0.001 |
| **Heart failure [HF]** | | | | |
| Prevalent |  |  |  | <0.001 |
| 0 | 5,141 (97%) | 2,176 (96%) | 2,965 (98%) |  |
| 1 | 170 (3.2%) | 100 (4.4%) | 70 (2.3%) |  |
| Incident |  |  |  | <0.001 |
| 0 | 4,791 (90%) | 1,992 (88%) | 2,799 (92%) |  |
| 1 | 520 (9.8%) | 284 (12%) | 236 (7.8%) |  |
| Follow-Up | 5.41 (4.68-6.42) | 5.28 (4.41-6.25) | 5.51 (4.81-6.51) | <0.001 |
| **Stroke** | | | | |
| Prevalent |  |  |  | <0.001 |
| 0 | 4,854 (91%) | 2,043 (90%) | 2,811 (93%) |  |
| 1 | 457 (8.6%) | 233 (10%) | 224 (7.4%) |  |
| Incident |  |  |  | 0.039 |
| 0 | 4,707 (89%) | 1,993 (88%) | 2,714 (89%) |  |
| 1 | 604 (11%) | 283 (12%) | 321 (11%) |  |
| Follow-Up | 9.7 (6.4-10.8) | 9.3 (5.4-10.5) | 9.8 (7.3-10.9) | <0.001 |
| **Type 2 diabetes [TD]** | | | | |
| Prevalent | 647 (12%) | 361 (16%) | 286 (9.3%) | <0.001 |
| Incident | 376 (14%) | 201 (18%) | 175 (11%) | <0.001 |
| **Medication** | | | | |
| Statins |  |  |  | <0.001 |
| 0 | 4,144 (77%) | 1,643 (72%) | 2,501 (81%) |  |
| 1 | 1,220 (23%) | 651 (28%) | 569 (19%) |  |
| *^1^*Median (Q1-Q3); n (%) | | | | |
| *^2^*Wilcoxon rank sum test; Pearson's Chi-squared test | | | | |
