## Supplementary figures for "Systematic comparison of observational and Mendelian Randomization estimates for cardiometabolic proteomic signatures"

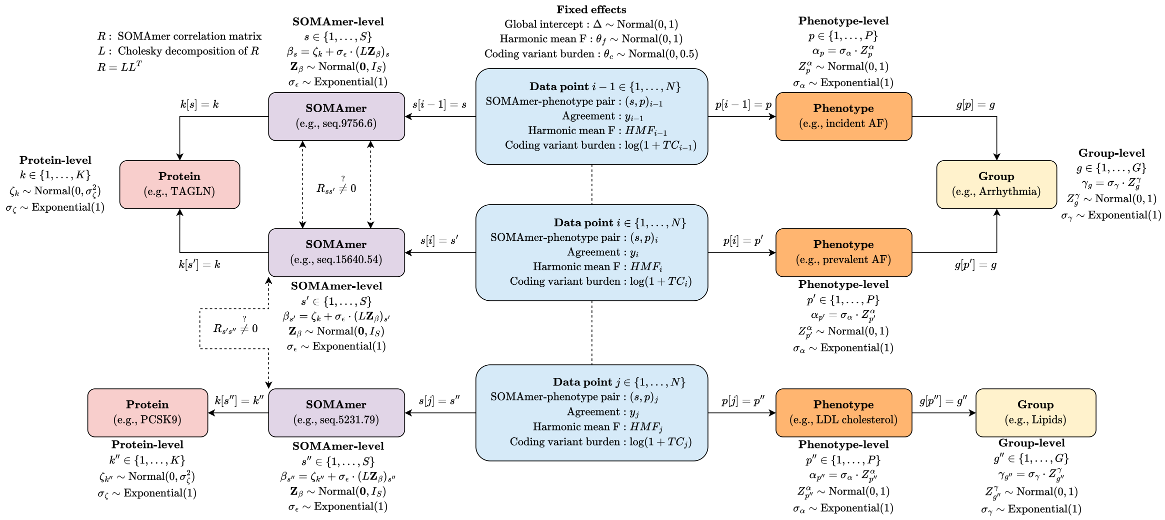


**Supplementary Figure 1: Structure of the hierarchical Bayesian model used to estimate directional agreement between observational and MR estimates.** Each data point $i$ corresponds to a SOMAmer-phenotype pair $(s, p)_i$, with binary agreement outcome $y_i$ and covariates: harmonic mean F-statistic ($HMF_i$) and total coding variant burden ($\log(1+TC_i)$). SOMAmers $s$ are linked to proteins $k$, which contribute to a protein-level random effect $\zeta_k$. Protein-level effects are shared across correlated SOMAmers via a multivariate normal prior with Cholesky factor $L$ derived from the SOMAmer correlation matrix $R$. Phenotypes $p$ are nested within phenotype groups $g$, contributing random effects $\alpha_p$ and $\gamma_g$, respectively. The fixed effects in the model include a global intercept $\Delta$, and coefficients for $HMF_i$ and coding variant burden. Dashed arrows indicate shared effects across SOMAmers, phenotypes, or proteins. Latent standard normal variables and exponential priors define the hierarchical prior distributions. This structure enables partial pooling across phenotype groups, phenotypes, proteins, and correlated SOMAmers while incorporating SNP quality and genetic architecture.

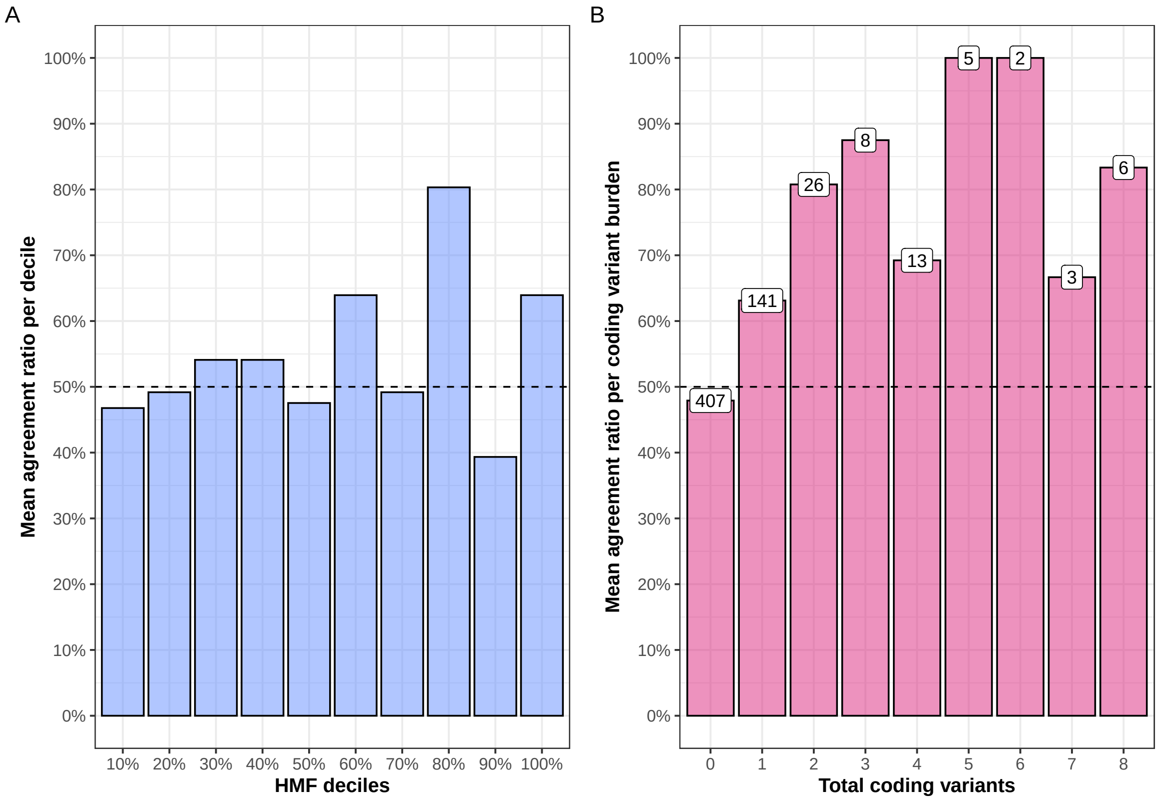


**Supplementary Figure 2: Observed agreement within covariates of Bayesian model. (A)** Observed directional agreement across HMF deciles. Each point represents the proportion of SOMAmer–phenotype pairs with matching effect directions between observational and MR estimates, stratified by decile of the harmonic mean F-statistic (HMF). No consistent trend was observed. **(B)** Observed directional agreement by coding variant burden. Agreement increases monotonically with the number of coding variants in the genetic instrument. Each point represents the proportion of SOMAmer–phenotype pairs with directional agreement, stratified by coding variant count.

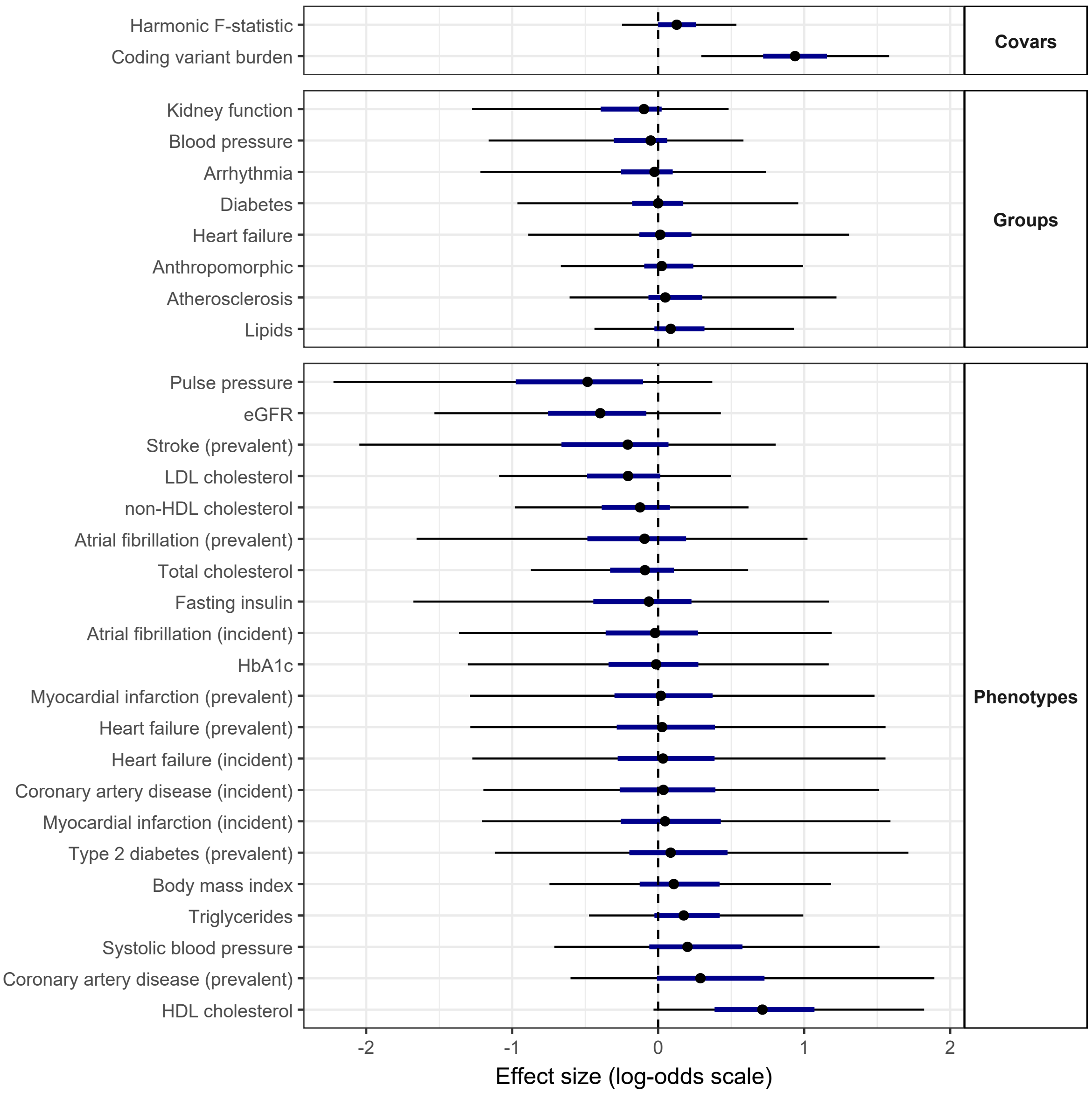


**Supplementary Figure 3: Posterior distributions of covariates.** Posterior medians (black circle) with 50% (dark blue) and 95% (black) credible intervals for all covariates, phenotype-group, and phenotype-specific effects from the hierarchical Bayesian logistic model. The dashed line marks no effect (log-odds = 0). Among all effects, the posterior for coding variant burden is the only one whose 95% credibility interval does not overlap zero.

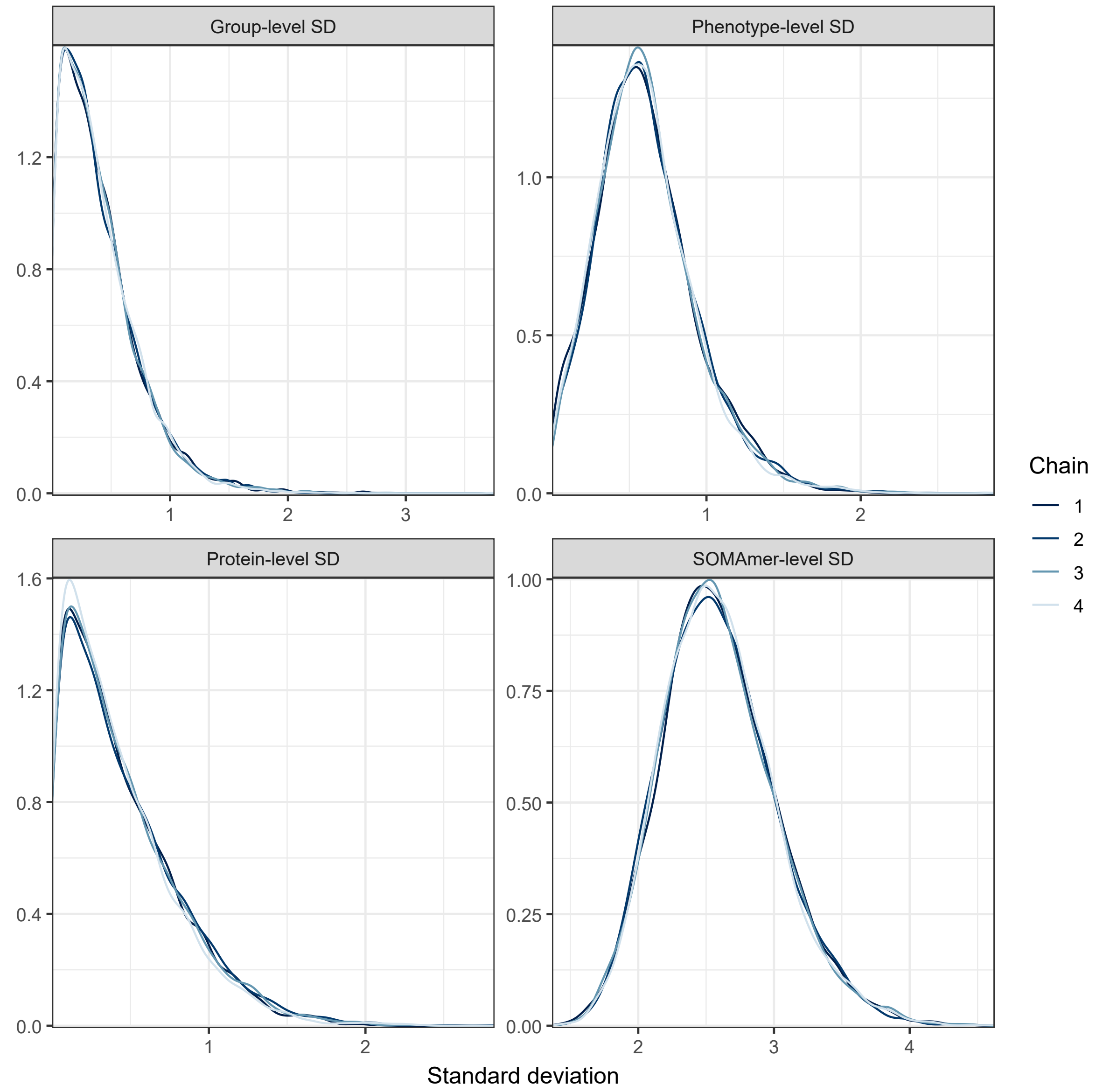


**Supplementary Figure 4: Posterior distributions of hierarchical standard deviations.** Posterior distributions for the group-, phenotype-, protein-, and SOMAmer-level standard deviations from the hierarchical Bayesian logistic model, estimated using four MCMC chains. The densities illustrate the posterior uncertainty in the degree of heterogeneity at each level of the model hierarchy.
